# High Brucellosis Prevalence and Risk Among School-Age Children in Kenyan Pastoral Health Facilities: A Facility-Based Surveillance Study

**DOI:** 10.64898/2026.03.09.26347949

**Authors:** Dismas C.O. Oketch, Ruth Njoroge, Isaac Ngere, John Gachohi, Walter Jaoko, Samuel Waiguru Muriuki, Athman Juma Mwatondo, Konongoi S. Limbaso, Mathew Muturi, Jodie Withall, John Mwaniki Njeru, Boru Ali, Boku Bodha, Lydia Kilowua, Nazaria Wanja Nyaga, Moshe Alando, David Maina, Samoel Ashimosi Khamadi, M. Kariuki Njenga, Roland T. Ashford, Eric M Osoro

**Affiliations:** Washington State University Global Health Program, Kenya, Nairobi 00100, Kenya; University of Nairobi, Department of Medical Microbiology and Immunology, Nairobi, Kenya; Washington State University, Paul G. Allen School for Global Health, Pullman, WA 99163, USA; Jomo Kenyatta University of Agriculture and Technology, School of Public Health, Nairobi, Kenya; University of Idaho, Department of Biological Sciences; Zoonotic Disease Unit, Government of Kenya, Nairobi, Kenya; Kenya Medical Research Institute, Centre for Virus Research, Nairobi, Kenya; Department of Veterinary Medicine, Dahlem Research School of Biomedical Sciences (DRS), Freie Universität Berlin, Berlin, Germany; Department of Bacteriology, Animal and Plant Health Agency (APHA), Weybridge, UK; Kenya Medical Research Institute, Centre for Microbiology Research, Nairobi, Kenya; County Government of Marsabit, Marsabit, Kenya; County Government of Kajiado, Kajiado, Kenya

**Keywords:** Brucellosis, Pastoralists, Zoonoses, Acute febrile illness, Kenya, Surveillance

## Abstract

**Background:** Brucellosis remains endemic in pastoral sub-Saharan Africa, yet systematic surveillance data on human prevalence and risk factors in these communities are limited. We estimated brucellosis prevalence and identified associated risk factors among febrile patients in two Kenyan pastoralist health facilities.

**Methodology/Principal Findings:** Prospective facility-based surveillance was conducted between February 2023 and July 2024 at Laisamis Sub-County Referral Hospital (Marsabit County) and Mailwa Health Center (Kajiado County). Consecutive enrollment included patients ≥1 year of age, with acute febrile illness and brucellosis-compatible symptoms. Venous blood samples underwent Rose Bengal Test (RBT) serology, real-time PCR for *Brucella* DNA, and indirect ELISA. Cases were defined using hierarchical laboratory criteria: RBT ≥1:8, PCR positivity among low-titer RBT samples, or combined low-titer RBT and ELISA positivity. Modified Poisson regression with robust variance estimation identified factors independently associated with brucellosis.

Among 441 enrolled participants, 67 (15.2%; 95% CI: 12.1%–18.8%) met brucellosis case criteria. Site-specific prevalence differed substantially: prevalence at Laisamis was 19.3% (95% CI: 15.4%–24.0%) compared to 3.5% at Mailwa (95% CI: 1.4%–8.6%; p<0.001). School-age children (5–14 years) experienced the highest age-specific prevalence at 31.0%, representing 38.8% of all cases despite comprising only 19.0% of the study population. In multivariable analysis, three factors remained independently associated with brucellosis: school-age group (5–14 years) compared to adults aged 15–49 years (adjusted prevalence ratio [aPR]: 2.42; 95% CI: 1.54–3.81; p<0.001), prolonged fever >7 days (aPR: 1.82; 95% CI: 1.19–2.77; p=0.005), and muscle pain (aPR: 2.44; 95% CI: 1.10–5.42; p=0.028). Sensitivity analysis restricting case classification to RBT ≥1:8 or PCR positivity (excluding low-titer RBT with ELISA+ cases) yielded associations of similar direction and magnitude.

**Conclusions/Significance:** Brucellosis prevalence in Kenyan pastoral health facilities is high, with school-age children emerging as a previously underrecognized high-risk group. Geographic variation in burden appears to align with ecological differences in livestock management and mobility. These findings suggest the need for age-targeted prevention strategies and diagnostic screening for prolonged fever in children. Coordinated action between clinical and veterinary sectors, aligned with Kenya’s national brucellosis control strategy, is needed to translate epidemiologic evidence into effective control efforts in pastoral communities.

**Author Summary:** Brucellosis is a bacterial infection spread from livestock to people, common in communities that depend on animals for their livelihood. In Kenya’s pastoral regions, families live in close contact with cattle, goats, sheep, and camels, regularly consuming fresh milk and assisting with animal births–activities that can transmit the disease. We tested 441 patients with fever at two health facilities in contrasting pastoral areas of Kenya and found that about 1 in 6 had brucellosis. The burden was not evenly distributed: one site had five times more cases than the other, likely reflecting differences in how livestock are managed and how much animals move and mix. Most unexpectedly, school-age children (5–14 years) had the highest rates of infection–nearly one in three tested positive–and accounted for almost 40% of all cases. These findings challenge the common assumption that brucellosis primarily affects adults through occupational exposure, and points to household milk-handling practices and children’s involvement in animal care as important but overlooked transmission routes. Patients with fever lasting more than a week were also at higher risk, suggesting that prolonged fever in children should prompt clinicians in pastoral areas to consider brucellosis. These findings highlight the need for prevention strategies that specifically address children’s exposures and for coordination between human health and veterinary services to reduce transmission at its source.

## Introduction

Brucellosis remains a leading zoonotic infection globally, with an estimated 2.1 million new human cases annually, though true incidence is substantially underreported in endemic regions [1]. Sub-Saharan Africa bears a disproportionate share of the burden, with community-based seroprevalence ranging from 2% to 44% depending on population and setting [2–4]. In pastoralist communities, close human–livestock contact increases transmission risk through multiple pathways: ingestion of unpasteurized dairy products, direct animal contact during milking, herding, or feeding, and occupational exposure during slaughter or birthing assistance. Beyond its direct health impacts, brucellosis also imposes major economic costs due to livestock infertility, trade restrictions, and reduced market opportunities [5].

In Kenya’s arid and semi-arid lands–which comprise 80% of the national territory and support approximately 40% of the population–pastoralist communities maintain daily contact with multiple livestock species (cattle, goats, sheep, and camels) through herding, milking, animal husbandry, and reproductive management tasks [6]. This human–animal interface supports transmission through herd-level conditions that sustain infection in livestock–including shared water points, inter-herd mixing during seasonal migration, and limited veterinary services–as well as direct human exposures such as milking, handling birthing materials, and consumption of unpasteurized dairy products including camel milk [7,8]. Despite endemic livestock brucellosis throughout the region, systematic surveillance for human brucellosis prevalence and transmission risk factors in pastoral populations remains limited.

Clinical diagnosis of brucellosis is complicated by non-specific symptoms overlapping malaria and typhoid fever, ranging from acute febrile illness to chronic musculoskeletal and neurological sequelae [9,10]. Accurate diagnosis requires multi-method approaches: the Rose Bengal Test (RBT) offers rapid screening but remains underutilized in sub-Saharan Africa; ELISA may improve specificity but depends on laboratory infrastructure; and real-time PCR provides highly specific detection of Brucella DNA but is constrained by laboratory capacity in resource-limited settings [9,11]. These constraints highlight the need for pragmatic, integrated diagnostics adapted to resource-limited settings.

Surveillance for brucellosis in sub-Saharan Africa is hindered by limited capacity for gold-standard methods like bacterial culture or paired serology, which are rarely feasible in pastoral settings due to biosafety concerns and difficulties in follow-up among mobile populations [12,13]. As a result, prevalence estimates vary widely, even within similar populations, and facility-based studies often rely on single diagnostic methods that underestimate the true burden [11,14]. In addition, prior studies have disproportionately focused on adults and older children presumed to have occupational exposure, overlooking household transmission pathways and age-specific vulnerabilities. Children’s roles in milk collection and animal care are rarely assessed, yet data from similar contexts suggest meaningful exposure risk [15,16]. A comprehensive understanding of demographic, clinical, and geographic drivers of infection remains lacking, limiting the design of effective interventions.

To address these gaps and inform locally adapted prevention strategies, we conducted prospective, facility-based surveillance embedded within a broader One Health investigation of brucellosis transmission dynamics in pastoral Kenya [17]. Our objectives were to: (1) estimate brucellosis prevalence among patients presenting with acute febrile illness at two ecologically distinct pastoral health facilities; (2) identify demographic, clinical, and exposure factors independently associated with brucellosis; and (3) assess geographic heterogeneity in disease burden and risk patterns.

## Methods

### Study Design and Setting

From February 2023 to July 2024, we conducted prospective surveillance at two sentinel health facilities: Laisamis Sub-County Referral Hospital (Marsabit County) and Mailwa Health Center (Kajiado County) (Fig 1). These sites were purposively selected based on reported livestock brucellosis burden and distinct ecological characteristics. Laisamis serves a highly mobile pastoralist population managing mixed herds of camels, cattle, sheep and goats across vast, communal rangelands. In contrast, Mailwa serves agropastoral communities where land tenure changes have led to subdivision of traditional grazing areas and more restricted livestock mobility [18,19]. This ecological heterogeneity created natural variation in transmission-relevant factors–including herd mixing, animal density, and mobility–enabling exploration of whether geographic variation in human brucellosis burden reflects local livestock management and environmental conditions. The study represents the human surveillance arm of a broader One Health longitudinal cohort examining *Brucella spp.* transmission dynamics in pastoral Kenya [17].

**Figure 1.**
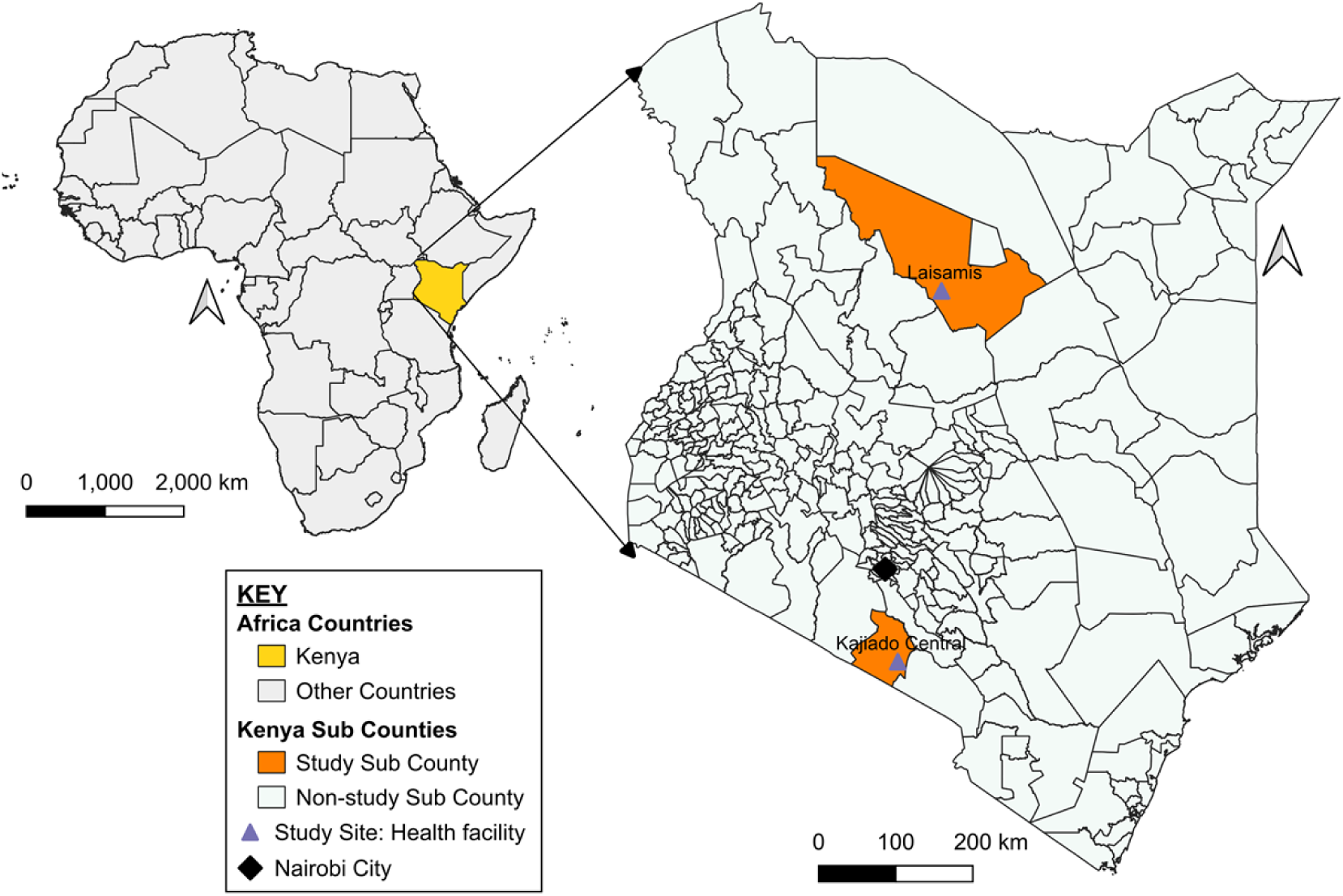
Map of the study sites. Orange areas indicate study subcounties; triangles show health facility locations; black diamond indicates capital city

### Study Population

Eligible participants were aged ≥1 year and presented with acute febrile illness (fever within 21 days or axillary temperature ≥38.0°C) accompanied by at least one brucellosis-compatible symptom: night sweats, joint pain, headache, fatigue, or anorexia. We excluded individuals with documented bleeding disorders precluding safe phlebotomy or those requiring immediate clinical stabilization. Enrollment proceeded among all consecutive patients meeting criteria during facility operating hours (Monday–Friday, 08:00–17:00).

### Data Collection

Trained study personnel administered structured questionnaires capturing demographic characteristics (age, sex, education, occupation, household composition, livestock holdings), clinical presentation (fever duration, symptom progression, prior treatments), and exposure history during the three months preceding illness onset. Exposure assessment focused on established transmission pathways: consumption of unpasteurized dairy products by animal species, direct contact with livestock (feeding, milking, herding), and participation in high-risk reproductive practices (assisting births, handling placental and fetal materials, managing abortion events). Species-specific frequencies and behaviors were recorded to enable construction of composite exposure variables reflecting raw milk consumption intensity (categorized as low/none, moderate, high) and animal contact diversity (≤1, 2, ≥3 species). All data were electronically captured in REDCap [20] with real-time validation, and field supervisors conducted regular audits with discrepancy resolution through source document review and participant re-contact.

### Laboratory Procedures

Venous blood samples (2–5 mL) were collected and transported under cold chain conditions to the field laboratory for serum separation, then shipped to Kenya Medical Research Institute Centre for Virus Research (KEMRI-CVR) for storage at −80°C until analysis. Rose Bengal Test (RBT) was performed using a semiquantitative serial dilution protocol, in which sera were tested at two-fold dilutions (neat through 1:32) against Rose Bengal antigen (Pourquier, IDEXX). This approach, which extends the standard qualitative RBT to provide titer information, has been validated in East African pastoral populations [11,21]. Titers ≥1:8 were considered indicative of infection, in line with prior findings including from East Africa [21]. Real-time PCR was performed using assays targeting the *Brucella* spp. IS*711* and *bcsp*31 genomic regions, respectively [22,23]. Samples were considered PCR positive when amplification was observed in both assay targets [21,22]. IgG antibodies were measured using a commercial indirect ELISA (Brucella IgG ELISA, IBL, Minneapolis, MN, USA) per manufacturer protocol, with classification based on lot-specific OD cutoffs.

### Brucellosis case classification

All enrolled participants met the acute febrile illness eligibility criterion and reported at least one brucellosis-compatible symptom, as defined above. Within this clinically eligible population, brucellosis cases were identified using a hierarchical, mutually exclusive analytic classification based on laboratory results:

1. RBT titer ≥1:8;
2. PCR detection of Brucella DNA with low-titer RBT reactivity (excluding participants meeting criterion 1); or
3. Low-titer RBT reactivity with ELISA IgG positivity (excluding participants meeting criteria 1 or 2).

Low-titer RBT reactivity was defined as visible agglutination at 1:2 or 1:4 dilution but negative at ≥1:8. Participants meeting any criterion were classified as cases; participants with complete laboratory results meeting none were classified as non-cases.

The requirement for RBT reactivity as a gating criterion for Tier 3 classification reflects evidence that RBT demonstrates sensitivity of 87.5–100% for detecting acute brucellosis in studies using appropriate comparison groups [24]. Low-titer RBT reactivity, while below the ≥1:8 threshold used for Tier 1, nonetheless indicates detectable agglutinating antibody whose diagnostic significance is strengthened when corroborated by a second serological method. ELISA IgG positivity without any RBT reactivity was not classified as a case, as isolated ELISA positivity may reflect cross-reactive antibodies or past resolved infection rather than current brucellosis [11,24]. This analytic classification was developed for epidemiologic inference in a facility-based surveillance context where culture and paired serology were not feasible. It differs from WHO confirmed case criteria, which require bacteriological confirmation or seroconversion. A sensitivity analysis restricting case classification to RBT ≥1:8 or PCR positivity was conducted to assess robustness to classification choice.

### Statistical Analysis

The study design, surveillance procedures, and laboratory methods were specified in the study protocol and have been described in detail [17]. Because bacterial culture and paired serology were not feasible in this surveillance context, an analytic case classification integrating RBT, PCR, and ELISA results was applied for epidemiologic analysis. Sensitivity analyses restricting the definition to RBT ≥1:8 or PCR positivity were conducted to evaluate the robustness of findings to case classification. Analyses were performed in R version 4.3.2 (R Core Team, 2023). Categorical variables are presented as frequencies and percentages; continuous variables as medians and interquartile ranges due to expected non-normality. Between-group comparisons employed chi-square or Fisher’s exact tests (when expected counts <5) for categorical variables, and Mann-Whitney U tests for continuous variables. Missingness across analytic variables was minimal and analyses were conducted using complete-case observations.

Brucellosis prevalence was estimated as the proportion of cases meeting laboratory criteria, with 95% confidence intervals calculated using the Wilson score method. Site-specific and age-stratified prevalence estimates were compared using two-proportion z-tests. Bivariate associations between case status and demographic, clinical, or exposure variables were examined using chi-square, Fisher’s exact, or Mann-Whitney U tests as appropriate. Composite exposure variables were constructed as follows: raw milk consumption intensity (low/none, moderate [single species or infrequent], or high [multiple species with frequent unpasteurized consumption]); animal contact diversity (low [≤1 species], moderate [2 species], or high [≥3 species]); and reproductive exposure risk (low, moderate, or high based on frequency of assisting animal births and handling placental or fetal materials).

Given overall outcome prevalence of 15.2% (>10%), modified Poisson regression with robust variance estimation was used for multivariable analysis to obtain unbiased prevalence ratios [25]. Variables achieving p<0.20 in bivariate analyses, along with *a priori* confounders (age group and sex), were entered into a full model. Age categories used in analysis reflected exposure patterns relevant to pastoral settings: <5 years (pre-school), 5–14 years (school-age), 15–49 years (working-age adults), and ≥50 years (older adults). The final model was developed through backward stepwise selection guided by Akaike Information Criterion. Model diagnostics assessed multi-colinearity using variance inflation factors and evaluated overdispersion using deviance-to-degrees-of-freedom ratios. Statistical significance was assessed at α=0.05 (two-sided).

Diagnostic stability was evaluated by repeating bivariate and multivariable analyses under a restricted analytic classification that excluded Tier 3 cases (low-titer RBT with ELISA IgG positivity). The restricted classification retained only RBT ≥1:8 or PCR positivity, thereby excluding 16 cases (all from Laisamis). This assessed whether core epidemiologic associations remained robust when applying more stringent laboratory criteria. Bivariate and multivariable analyses were repeated identically with the restricted classification. Associations were classified as stable if they maintained consistent direction, overlapping 95% confidence intervals, and statistical significance (α=0.05) across both case classification.

### Ethical Considerations

Written informed consent was obtained from participants ≥18 years; those <18 years provided parental/guardian consent and assent as appropriate. The study was approved by KEMRI Scientific and Ethics Review Unit (Protocol No. 4405), reliance obtained from the Washington State University Institutional Review Board, and authorized by the Kenya Ministry of Health and National Commission for Science, Technology and Innovation (License No: NACOSTI/P/22/17621).

## Results

### Participant Enrolment and Study Population

Of 1,743 individuals with acute febrile illness screened, 1,085 (62.3%) did not meet eligibility criteria. Of 658 eligible participants, 215 (32.7%) were not enrolled. Enrollment was conducted during routine facility operating hours (Monday–Friday, 08:00–17:00). Non-enrollment primarily reflected operational constraints such as limited staffing capacity and participant time limitations rather than systematic exclusion based on clinical characteristics. Following enrollment of 443 participants, 2 (0.5%) were excluded due to incomplete laboratory data, yielding a final analytic sample of 441 participants (441/658 = 67.0% of eligible individuals). Participant flow is presented in Fig 2.

**Fig 2.**
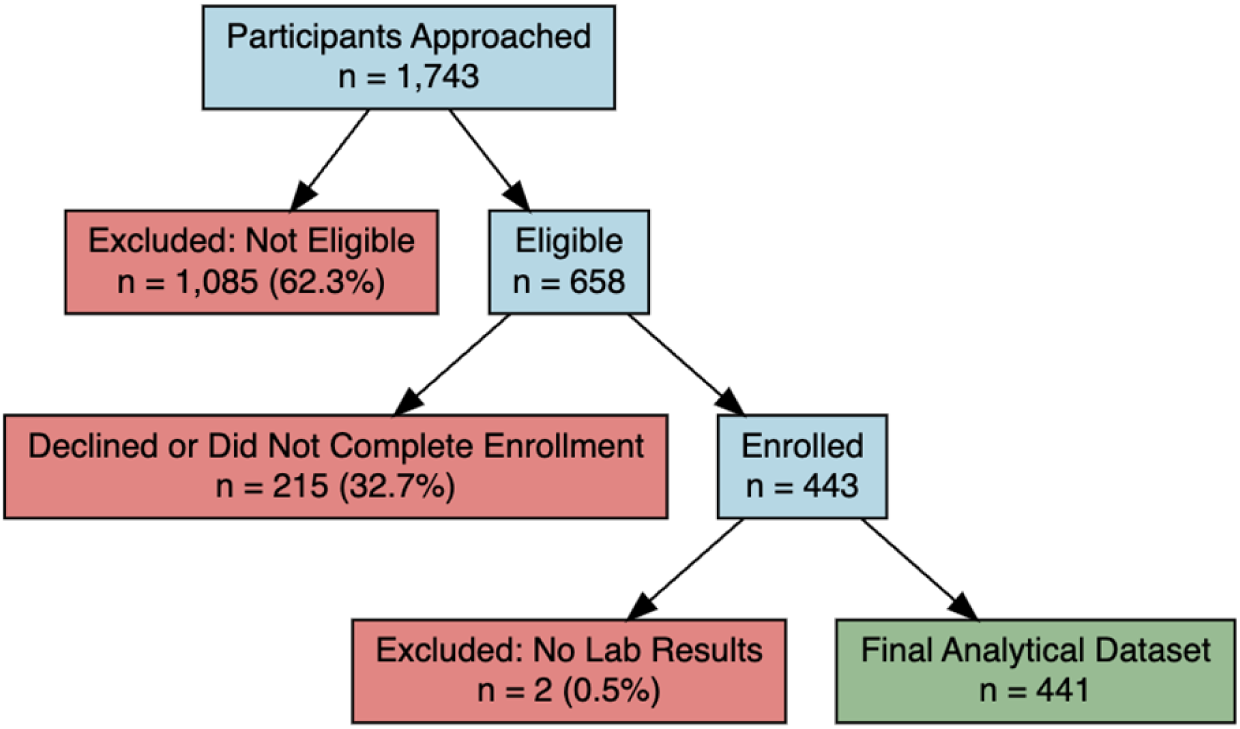
Participant enrolment and exclusion flowchart for brucellosis surveillance study across two pastoral sites in Kenya, February 2023-July 2024

### Brucellosis Prevalence

Among 441 participants, 67 (15.2%; 95% CI: 12.1%–18.8%) met brucellosis case criteria (Fig 3). The hierarchical classification identified 26 cases (38.8%) via RBT titer ≥1:8 (Tier 1), 25 (37.3%) via PCR positivity with low-titer RBT reactivity (Tier 2), and 16 (23.9%) via low-titer RBT reactivity with ELISA IgG positivity (Tier 3). The detailed diagnostic patterns are presented in Table S1.

**Fig 3.**
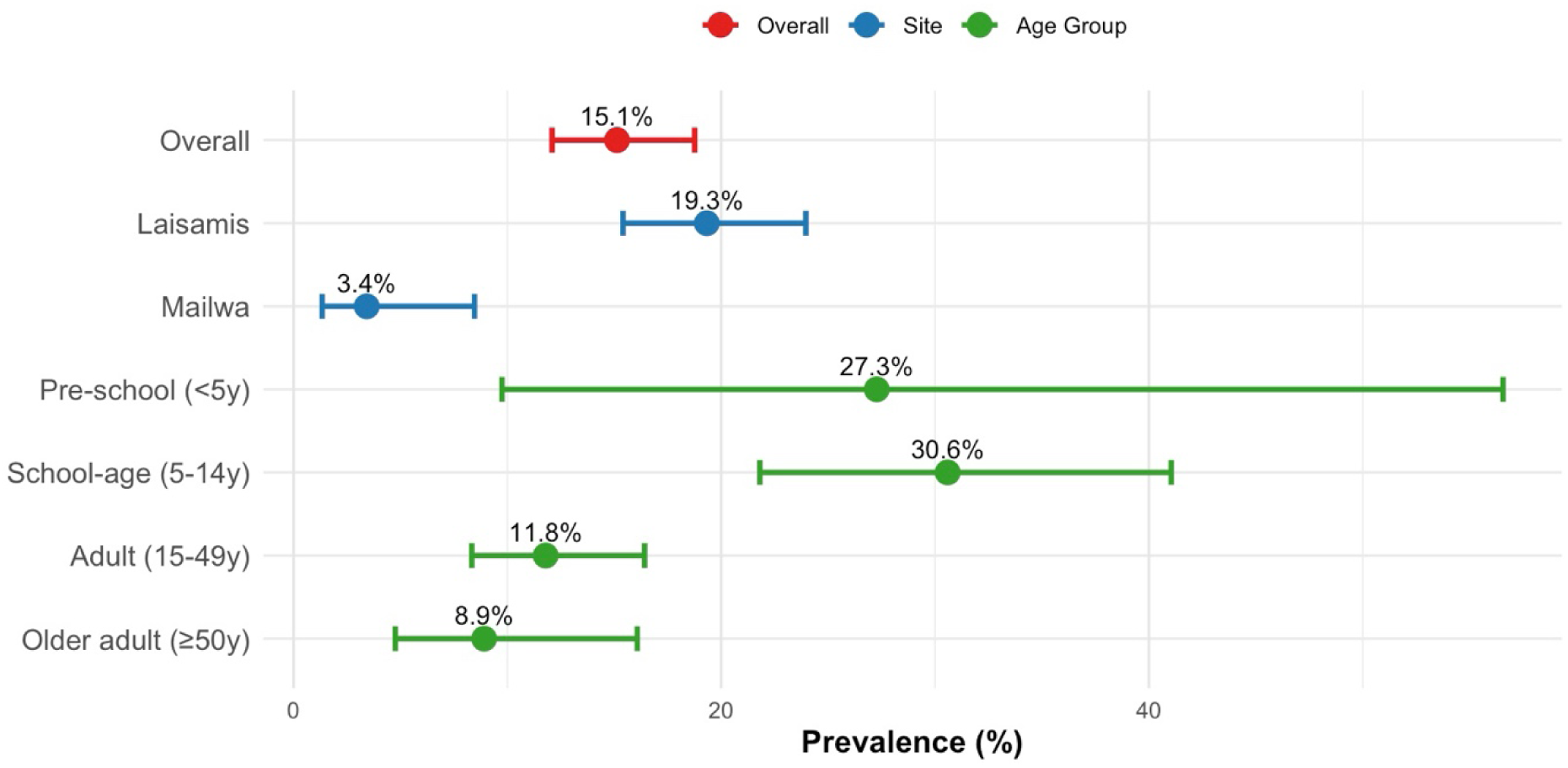
Brucellosis prevalence with 95% confidence intervals by study site and age group among febrile patients in pastoral Kenya. School-age children (5–14 years) demonstrated the highest prevalence across age strata

Geographic variation was substantial: Laisamis reported 19.3% prevalence (95% CI: 15.4%–24.0%) compared to 3.5% in Mailwa (95% CI: 1.4%–8.6%; p<0.001), representing a 5.5-fold difference between sites (Fig 3). Age-stratified prevalence estimates revealed marked variation across age groups. School-age children (5–14 years) experienced the highest prevalence at 31.0%, followed by pre-school children (<5 years) at 27.3%. In contrast, adults aged 15–49 years reported 11.8% prevalence, and older adults (≥50 years) 8.9%.

### Sociodemographic, Clinical, and Exposure Characteristics

Cases were substantially younger than non-cases: median age 18.0 years (IQR: 11.0–34.7) versus 34.0 years (IQR: 20.0–49.0; p<0.001). School-age children represented 38.8% of cases but only 19.0% of the total population. Males comprised 59.7% of cases compared to 39.8% of non-cases (p=0.003). Geographically, 94.0% of cases occurred at Laisamis. Bivariate comparisons of demographic, clinical, and exposure characteristics are presented in Table 1.

**Table 1.**
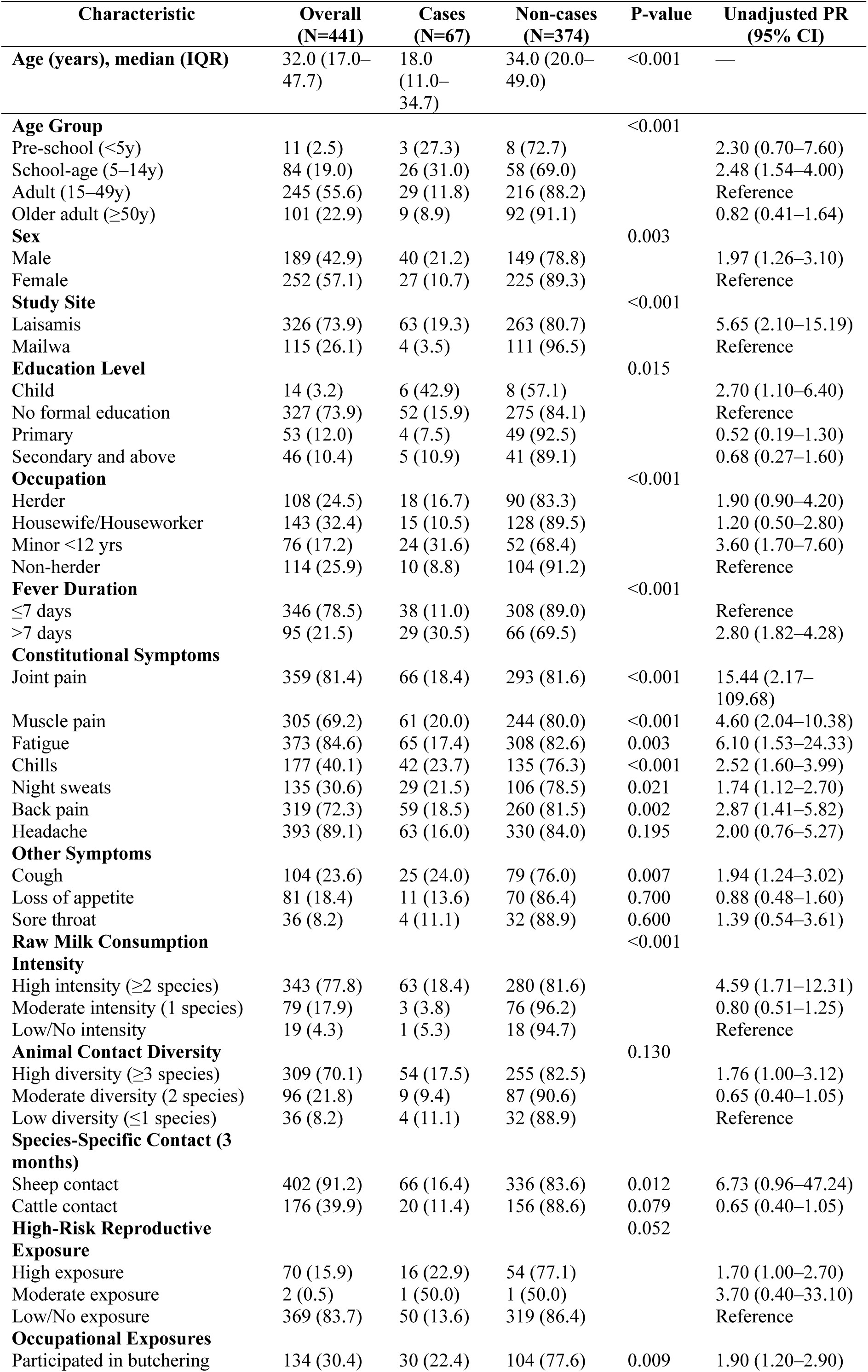
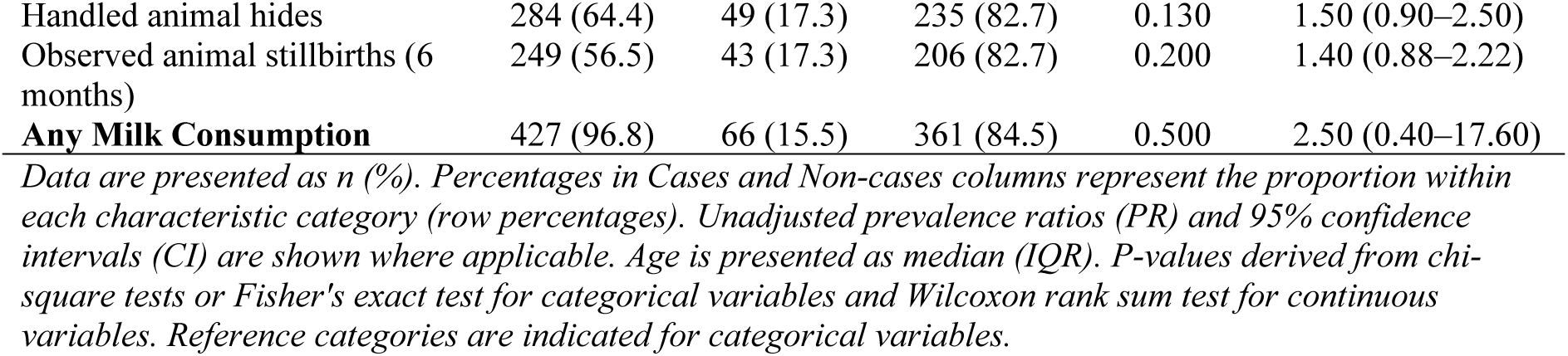
Sociodemographic, Clinical, and Exposure Characteristics by Brucellosis Case Status.

Cases presented with different clinical profiles from non-cases. Prolonged fever (>7 days) was twice as common in cases (43.3% vs. 17.6%; p<0.001), while constitutional symptoms were near-universal among cases: joint pain (98.5% vs. 77.8%; p<0.001), muscle pain (91.0% vs. 65.2%; p<0.001), fatigue (97.0% vs. 81.8%; p=0.003), and chills (62.7% vs. 36.1%; p<0.001). In addition, respiratory and musculoskeletal manifestations–back pain (88.1% vs. 69.1%; p=0.002), cough (37.3% vs. 21.0%; p=0.007)–were more frequent in cases (Table 1).

Exposure patterns differed between cases and non-cases. Raw milk consumption was almost universal in both groups (96.8% and 96.0%, respectively), but consumption intensity was different: 94.0% of cases reported high-intensity consumption (multiple livestock species) versus 74.9% of non-cases (p<0.001). Contact with small ruminants was also nearly ubiquitous in cases: 98.5% reported recent sheep contact compared to 89.4% of non-cases (p=0.012). Participation in high-risk occupational practices–butchering, handling reproductive materials–occurred in 44.8% of cases versus 27.8% of non-cases (p=0.009).

### Multivariable Analysis

In modified Poisson regression, three factors independently predicted brucellosis (Fig 4). School-age children (5–14 years) demonstrated the strongest association with brucellosis risk (adjusted prevalence ratio [aPR]: 2.42; 95% CI: 1.54–3.81; p<0.001) compared to adults aged 15–49 years. Prolonged fever exceeding 7 days almost doubled the risk (aPR: 1.82; 95% CI: 1.19–2.77; p=0.005), while muscle pain independently increased risk (aPR: 2.44; 95% CI: 1.10–5.42; p=0.028). Of note, pre-school age (<5 years) approached significance with similar magnitude of effect (aPR: 2.33; 95% CI: 1.00–5.42; p=0.050). Joint pain and recent sheep contact, though showing higher point estimates, did not reach statistical significance in the adjusted model (aPR: 5.30 and 4.45, respectively). Older adults (≥50 years) showed no significant elevation in risk (aPR: 0.65; 95% CI: 0.32–1.30; p=0.22).

**Figure 4.**
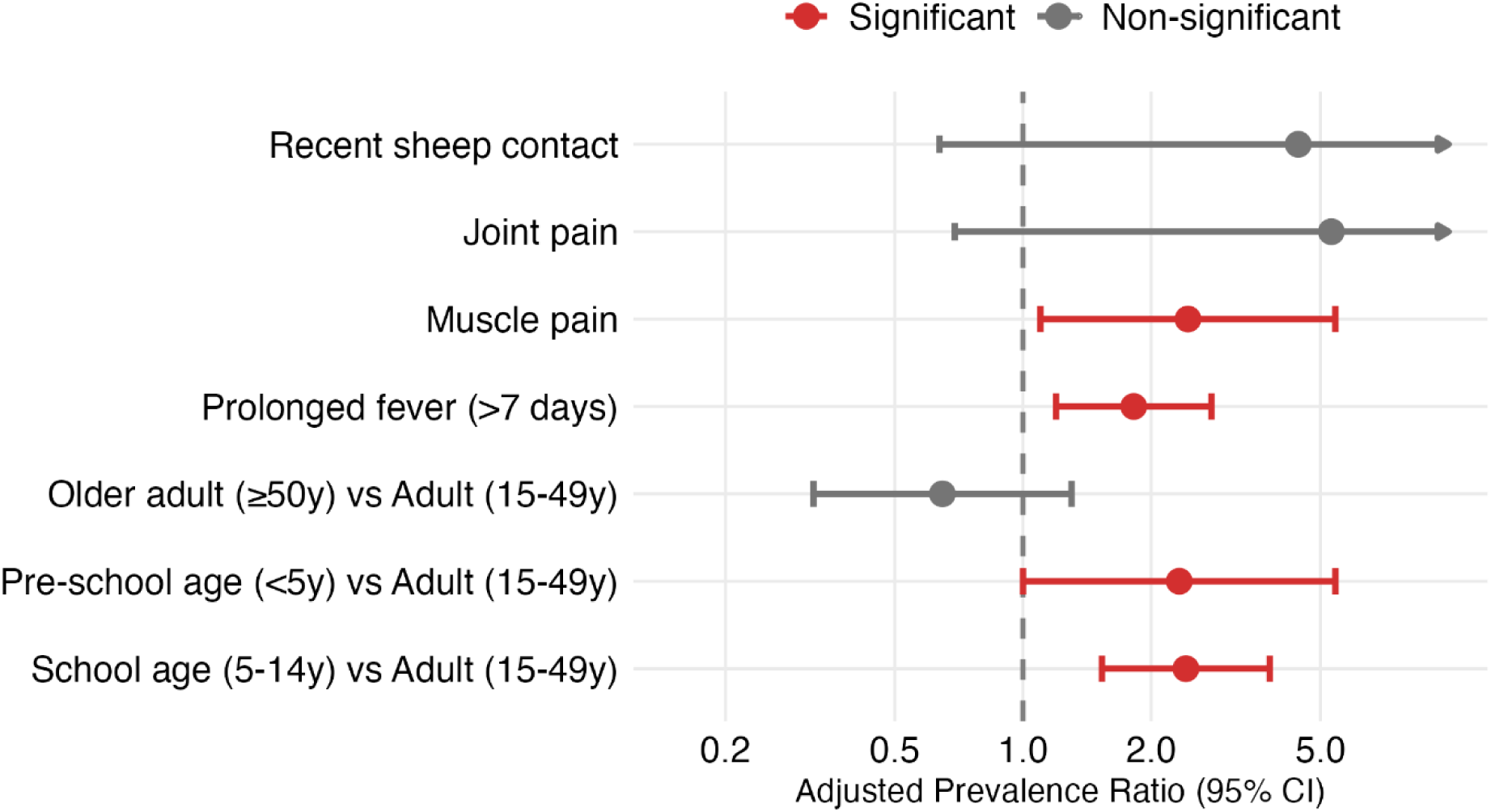
Multivariable risk factors for brucellosis seropositivity among patients with acute febrile illness, pastoral Kenya. Adjusted prevalence ratios (aPR) and 95% confidence intervals (95% CI) estimated using modified Poisson regression with robust variance. The upper 95% CIs for joint pain (40.7) and recent sheep contact (30.9) exceed the x-axis scale and are truncated with arrows for clarity.

### Sensitivity Analysis

To assess diagnostic stability, we repeated multivariable analysis using a restricted set of case definitions (RBT ≥1:8 or PCR+), excluding the 16 cases with low-titer RBT+/ELISA+ serology. This restriction reduced overall prevalence from 15.2% to 11.6%. Laisamis prevalence declined from 19.3% to 14.5% (all 16 borderline cases occurred at this site), while Mailwa remained unchanged at 3.5%. Age-stratified analysis showed differential case loss: older adults lost 33% of cases, whereas school-age children lost only 15% (Table S2)

Despite this diagnostic restriction, core associations remained robust and directionally consistent (Table S3). School-age children retained the strongest association (aPR: 2.95; 95% CI: 1.72–5.06; p<0.001), representing a 22% increase in the point estimate compared to the primary model (2.41 to 2.95). Prolonged fever remained significant (aPR: 2.10; 95% CI: 1.28–3.43; p=0.003), representing a 15% increase in the point estimate (1.82 to 2.10). Pre-school age reached statistical significance (aPR: 3.29; 95% CI: 1.36–7.96; p=0.008), representing a 41% increase in the point estimate (2.33 to 3.29). Muscle pain lost statistical significance (aPR: 2.12; 95% CI: 0.85–5.26; p=0.106), representing a 13% decrease in the point estimate (2.44 to 2.12).

## Discussion

This study demonstrates a substantial burden of brucellosis among febrile patients attending pastoral health facilities in Kenya, with an overall prevalence of 15.2%, and identifies school-age children as a previously underrecognized high-risk group. This prevalence exceeds most prior facility-based estimates in East Africa (5–13%) [14,16,26]. The higher prevalence likely reflects two complementary factors: the integrated diagnostic approach (RBT, PCR, ELISA) combined serologic and molecular tests to detect both acute and chronic infections; and persistent transmission-enabling conditions–limited livestock vaccination, minimal milk pasteurization, and reliance on fresh dairy products for household nutrition [11,27]

A notable finding was the concentration of brucellosis among school-age children (31.0% prevalence; 38.8% of all cases), departing from typical occupational risk patterns in pastoral epidemiology. While similar patterns have been reported in neighboring Tanzania [16], this contrasts with most East African studies emphasizing adult occupational risk [14,28]. Several mechanisms may explain this vulnerability pattern. Cultural and dietary practices often prioritize unpasteurized milk for children, and children frequently participate in animal husbandry activities such as feeding, watering, and caring for small ruminants. Differences in cumulative exposure history and immunologic experience between children and adults may also contribute, although evidence for protective immunity from repeated exposure remains limited [20]. This age-concentrated burden suggests prevention strategies must address household-level milk-handling practices and children’s participation in animal care.

The five-fold geographic difference in prevalence (19.3% Laisamis vs. 3.5% Mailwa) reflects distinct ecological systems. Laisamis, characterized by extensive pastoral mobility and seasonal herd migration, may facilitate more frequent inter-herd contact and *Brucella* spp. transmission. Mailwa has undergone land tenure restructuring with grazing subdivision and settlement, which may constrain livestock movement. This ecological distinction aligns with spatial mapping of brucellosis risk in Kenya, where mobile pastoralism is associated with higher livestock seroprevalence [3,18]. The geographic variation demonstrates that human brucellosis burden follows ecological determinants of herd-level disease dynamics and transmission intensity rather than uniform risk across pastoral regions, though the two-site design limits the ability to fully disentangle site-specific factors from broader ecological mechanisms. Interpretation of individual-level risk factors should therefore consider the possibility of residual confounding by site-level ecological differences.

Prolonged fever (>7 days) demonstrated robustness as an independent clinical predictor (aPR: 1.82; p=0.005), persisting across both primary and restricted case definitions (Table S3). This aligns with brucellosis’s classically subacute course, distinguishing it from acute infections like malaria or pneumonia that typically resolve within a few days [29]. In resource-limited pastoralist settings where laboratory confirmation may be delayed, prolonged fever–especially when accompanied by musculoskeletal symptoms–should raise clinical suspicion for brucellosis. Cough was also more frequently reported among cases (37.3% vs. 21.0%; p=0.007), but given its non-specificity in outpatient febrile illness, this finding should be interpreted cautiously. The association may reflect co-infections or symptom clustering rather than a diagnostically discriminating feature.

Muscle pain was highly prevalent among cases (91.0% vs. 65.2%) and significantly associated with case status in the primary analysis (p=0.028). This association attenuated in the restricted analysis (p=0.106), likely reflecting reduced statistical power with fewer cases (n=51) rather than absence of clinical relevance. Prolonged fever, by contrast, retained predictive value across models, reinforcing its utility as a consistent clinical signal. The co-occurrence of prolonged fever, musculoskeletal pain may help frontline clinicians differentiate brucellosis from other febrile illnesses in endemic pastoral areas.

Raw milk consumption and direct animal contact did not persist as independent risk factors in multivariable analysis. Several mechanisms may explain this finding. First, these exposures are near-universal in this population (96.8% reported any milk consumption; 90.7% reported sheep contact), limiting individual-level variation in adjusted models. Second, the high prevalence of these exposures across the study population limits their ability to discriminate individual-level risk in multivariable models. Third, collinearity or confounding by age, sex, or clinical presentation may have attenuated their independent effects. The independent predictors that emerged in adjusted analysis were school-age age group, prolonged fever, and muscle pain–suggesting that clinical and demographic factors discriminate cases more effectively than exposure history in this setting. Given strong site clustering of cases (94% from Laisamis), some exposure associations–particularly raw milk consumption intensity–may partially reflect site-level ecology and pastoral mobility patterns rather than individual-level causal effects. The multivariable model adjusted for site, but complete disentanglement of individual and ecological effects was not possible given the observed distribution.

The hierarchical case classification included 16 participants (23.9% of cases) identified through low-titer RBT reactivity with ELISA confirmation (Tier 3). These cases met laboratory criteria but relied on less stringent serologic evidence than Tiers 1–2. Sensitivity analyses excluding Tier 3 cases demonstrated that key associations remained consistent in direction and magnitude, supporting the robustness of findings and the epidemiologic validity of the primary analytic classification.

Prevention and surveillance strategies addressing school-age children require approaches distinct from occupational risk mitigation. Household-level interventions including improved milk handling and age-stratified education regarding animal contact represent potential complements to traditional pastoral health promotion. Health facility staff could systematically assess children with prolonged fever for brucellosis in high-prevalence sites. The diagnostic approach combining RBT, PCR, and ELISA demonstrated operational feasibility; facility-level capacity for RBT screening with referral pathways for confirmatory testing warrants consideration as a practical implementation pathway. Coordination between clinical and veterinary services, informed by shared laboratory results, enables linked surveillance across human and animal health sectors.

Several limitations warrant acknowledgment. The cross-sectional design precludes causal inference; prospective follow-up from the nested longitudinal cohort will help establish temporality. As a facility-based study, our prevalence estimates likely overestimate community-level burden by capturing symptomatic individuals; parallel community serosurveys will estimate community-level seroprevalence and asymptomatic infection burden. The hierarchical case definition included participants with low-titer RBT and ELISA positivity; however, sensitivity analysis excluding these individuals demonstrated stable associations, supporting their inclusion as brucellosis cases. The lack of systematic documentation for 215 non-enrolled eligible participants limits assessment of selection bias; if non-enrolees differed meaningfully in exposure or susceptibility, brucellosis prevalence could be biased. Finally, findings are based on two pastoralist populations and may not generalize to other ecological or occupational settings without caution.

This study identified a brucellosis prevalence of 15.2% among febrile patients in pastoral health facilities in Kenya, with marked geographic variation reflecting ecological differences in livestock management. School-age children emerged as a previously underrecognized risk population. These findings support the integration of age-sensitive surveillance and prevention strategies within Kenya’s national brucellosis control framework [30], and highlight the need to address household-level transmission pathways affecting children in pastoral communities. Coordinated action between clinical and veterinary sectors at the county level will be essential for translating epidemiologic evidence into effective disease control.

## Acknowledgments

We thank the study participants and their families for their participation. We acknowledge the clinical and administrative staff at Laisamis Sub-County Referral Hospital and Mailwa Health Center for their support during data collection and sample acquisition. Laboratory analyses were conducted at the Kenya Medical Research Institute Centre for Virus Research (KEMRI-CVR), with technical support from the Animal and Plant Health Agency (APHA), Weybridge, UK. We gratefully acknowledge the County Governments of Marsabit and Kajiado for facilitating field operations and providing epidemiological context. We thank the Kenya Zoonotic Disease Unit for collaboration and surveillance integration support.

## Author Contributions

Conceptualization: EMO, DCO, RTA, MKN. Methodology: EMO, DCO, JG, RTA, JW, SAK, KSL, RN, MM, LK, NWN. Data curation: RN, IN, KSL, BA, BB, SWM. Formal analysis: EMO, JG. Investigation: EMO, DCO, RN, IN, KSL. Laboratory work: RTA, JW, SWM, KSL, SAK, MA. Supervision: MKN, SAK, RTA. Writing – original draft: EMO. Writing – review and editing: EMO, DCO, JG, RTA, AJM, JMN, MKN. Administrative and field coordination: BA, BB. All authors reviewed and approved the final manuscript.

## Ethical Approval and Consent to Participate

Written informed consent was obtained from participants aged ≥18 years. Parental/guardian consent and assent were obtained for participants <18 years. The study was approved by the KEMRI Scientific and Ethics Review Unit (Protocol No. 4405), reliance obtained from Washington State University Institutional Review Board, and authorized by the Kenya Ministry of Health and National Commission for Science, Technology and Innovation (License No: NACOSTI/P/22/17621).

## Data Availability

De-identified individual-level data underlying the findings of this study are available upon reasonable request from the Washington State University Global Health Program and the Kenya Medical Research Institute for researchers who meet the criteria for access to confidential data. Data access may require institutional data sharing agreements and ethical approval where applicable.

## Supporting Information

S1 Table. Brucellosis case classification by diagnostic criteria and study site (n=441), February 2023–July 2024.

S2 Table. Sensitivity analysis of brucellosis prevalence by case definition and study stratum.

S3 Table. Adjusted prevalence ratios for brucellosis risk factors: primary versus restricted case definitions.

S4 Checklist. STROBE checklist for cross-sectional studies.

## References

1. Laine CG, Johnson VE, Scott HM, Arenas-Gamboa AM. Global Estimate of Human Brucellosis Incidence. Emerg Infect Dis. 2023;29: 1789. doi:10.3201/EID2909.230052

2. Njeru J, Melzer F, Wareth G, El-Adawy H, Henning K, Pletz MW, et al. Human Brucellosis in Febrile Patients Seeking Treatment at Remote Hospitals, Northeastern Kenya, 2014–2015. Emerg Infect Dis. 2016;22: 2160. doi:10.3201/EID2212.160285

3. Osoro EM, Munyua P, Omulo S, Ogola E, Ade F, Mbatha P, et al. Strong association between human and animal brucella seropositivity in a linked study in Kenya, 2012-2013. American Journal of Tropical Medicine and Hygiene. 2015;93: 224–231. doi:10.4269/ajtmh.15-0113

4. Asrie F, Birhan M, Dagnew M, Berhane N. Prevalence of human brucellosis in Ethiopia: systematic review and meta-analysis. BMC Infect Dis. 2025;25: 365. doi:10.1186/S12879-025-10502-8

5. Mcdermott JJ, Grace D, Zinsstag J. Economics of brucellosis impact and control in low-income countries. Rev Sci Tech. 2013. doi:10.20506/rst.32.1.2197

6. Kenya National Bureau of Statistics. 2019 Kenya Population and Housing Census. Kenya National Bureau of Statistics. Nairobi; 2019 Jul. Available: https://www.knbs.or.ke/?p=5621

7. McDermott JJ, Arimi SM. Brucellosis in sub-Saharan Africa: Epidemiology, control and impact. Vet Microbiol. 2002;90: 111–134. doi:10.1016/S0378-1135(02)00249-3

8. Corbel MMJ. Brucellosis in Humans and Animals. World Health Organization. World Health Organization, Food and Agriculture Organization of the United Nations, World Organization for Animal Health, Geneva, Switzerland; 2006. doi:10.2105/AJPH.30.3.299

9. Dean A, Crump L, Greter H, Hattendorf J, Schelling E, Zinsstag J. Clinical manifestations of human brucellosis: a systematic review and meta-analysis. PLoS Negl Trop Dis. 2012.

10. Köse Ş, Senger SS, Akkoçlu G, Kuzucu L, Ulu Y, Ersan G, et al. Clinical manifestations, complications, and treatment of brucellosis: Evaluation of 72 cases. Turk J Med Sci. 2014. doi:10.3906/sag-1112-34

11. Díaz R, Casanova A, Ariza J, Moriyón I. The Rose Bengal Test in Human Brucellosis: A Neglected Test for the Diagnosis of a Neglected Disease. PLoS Negl Trop Dis. 2011;5: e950. doi:10.1371/JOURNAL.PNTD.0000950

12. Franc KA, Krecek RC, Häsler BN, Arenas-Gamboa AM. Brucellosis remains a neglected disease in the developing world: A call for interdisciplinary action. BMC Public Health. 2018;18. doi:10.1186/S12889-017-5016-Y,

13. WHO Regional Office for Africa. Technical Guidelines for Integrated Disease Surveillance and Response in the African Region: Third edition | WHO | Regional Office for Africa. 2019 [cited 20 Oct 2025]. Available: https://www.afro.who.int/publications/technical-guidelines-integrated-disease-surveillance-and-response-african-region-third

14. Njeru J, Wareth G, Melzer F, Henning K, Pletz MW, Heller R, et al. Systematic review of brucellosis in Kenya: Disease frequency in humans and animals and risk factors for human infection. BMC Public Health. 2016;16. doi:10.1186/s12889-016-3532-9

15. Bett B, Jost C, Allport R, Mariner J. Using participatory epidemiological techniques to estimate the relative incidence and impact on livelihoods of livestock diseases amongst nomadic pastoralists in Turkana South District, Kenya. Prev Vet Med. 2009;90: 194–203. doi:10.1016/J.PREVETMED.2009.05.001

16. Bodenham RF, Lukambagire AHS, Ashford RT, Buza JJ, Cash-Goldwasser S, Crump JA, et al. Prevalence and speciation of brucellosis in febrile patients from a pastoralist community of Tanzania. Scientific Reports 2020 10:1. 2020;10: 1–11. doi:10.1038/s41598-020-62849-4

17. Oketch D, Njoroge R, Ngere I, Gachohi J, Waiguru S, Omia D, et al. Design of a Prospective Human–Animal Cohort Study to Evaluate the Role of Camels and Other Livestock Species in the Transmission of Brucella spp. to Humans in Kenya. Int J Environ Res Public Health. 2025;22: 1859. doi:10.3390/ijerph22121859

18. Akoko JM, Mwatondo A, Muturi M, Wambua L, Abkallo HM, Nyamota R, et al. Mapping brucellosis risk in Kenya and its implications for control strategies in sub-Saharan Africa. Sci Rep. 2023;13: 1–11. doi:10.1038/S41598-023-47628-1;SUBJMETA=1470,326,41,631,692,699;KWRD=DISEASES,INFECTIOUS-DISEASE+EPIDEMIOLOGY

19. Rotich SJ, Funder M, Marani M, Nathan I. Climate change adaptation and land tenure: exploring pastoral adaptation strategies under communal and private land ownership in Kenya. Local Environ. 2025;30: 37–57. doi:10.1080/13549839.2024.2428207;REQUESTEDJOURNAL:JOURNAL:CLOE20;SUBPAGE:STRING:ACCESS

20. Harris PA, Taylor R, Thielke R, Payne J, Gonzalez N, Conde JG. Research electronic data capture (REDCap)-A metadata-driven methodology and workflow process for providing translational research informatics support. J Biomed Inform. 2009. doi:10.1016/j.jbi.2008.08.010

21. Lukambagire AHS, Mendes ÂJ, Bodenham RF, McGiven JA, Mkenda NA, Mathew C, et al. Performance characteristics and costs of serological tests for brucellosis in a pastoralist community of northern Tanzania. Scientific Reports 2021 11:1. 2021;11: 1–11. doi:10.1038/s41598-021-82906-w

22. Probert WS, Schrader KN, Khuong NY, Bystrom SL, Graves MH. Real-time multiplex PCR assay for detection of Brucella spp., B. abortus, and B. melitensis. J Clin Microbiol. 2004;42: 1290–1293. doi:10.1128/JCM.42.3.1290-1293.2004

23. Matero P, Hemmilä H, Tomaso H, Piiparinen H, Rantakokko-Jalava K, Nuotio L, et al. Rapid field detection assays for Bacillus anthracis, Brucella spp., Francisella tularensis and Yersinia pestis. Clinical Microbiology and Infection. 2011;17: 34–43. doi:10.1111/j.1469-0691.2010.03178.x

24. Ekiri AB, Kilonzo C, Bird BH, Vanwormer E, Wolking DJ, Smith WA, et al. Utility of the Rose Bengal Test as a Point-of-Care Test for Human Brucellosis in Endemic African Settings: A Systematic Review. J Trop Med. 2020;2020: 6586182. doi:10.1155/2020/6586182

25. Zou G. A Modified Poisson Regression Approach to Prospective Studies with Binary Data. Am J Epidemiol. 2004;159: 702–706. doi:10.1093/AJE/KWH090

26. Sileshi B, Gizaw S, Merkeb B, Bekele T, Tadesse W, Kezali J, et al. Sero-prevalence of human brucellosis and associated factors among febrile patients attending Moyale Primary Hospital, Southern Ethiopia, 2023: Evidences from pastoralist community. PLoS Negl Trop Dis. 2024;18: e0012715. doi:10.1371/JOURNAL.PNTD.0012715

27. Loubet P, Magnan C, Salipante F, Pastre T, Keriel A, O’callaghan D, et al. Diagnosis of brucellosis: Combining tests to improve performance. PLoS Negl Trop Dis. 2024;2024-September. doi:10.1371/JOURNAL.PNTD.0012442,

28. Tumwine G, Matovu E, Kabasa JD, Owiny DO, Majalija S. Human brucellosis: sero-prevalence and associated risk factors in agro-pastoral communities of Kiboga District, Central Uganda. BMC Public Health. 2015;15: 1–8. doi:10.1186/S12889-015-2242-Z/TABLES/3

29. Dean AS, Crump L, Greter H, Schelling E, Zinsstag J. Global Burden of Human Brucellosis: A Systematic Review of Disease Frequency. PLoS Negl Trop Dis. 2012;6. doi:10.1371/JOURNAL.PNTD.0001865,

30. Zoonotic Disease Unit. National Strategy for the Prevention and Control of Brucellosis in Humans and Animals in Kenya (2021–2040). Nairobi; 2021.

